# Factors that lead to patient mistrust of the healthcare system in Hargeisa, Somaliland: a qualitative exploration

**DOI:** 10.1101/2023.11.23.23298819

**Authors:** Hersi Y Ahmed, Abdillahi A Mussa, Abdisamed M Yousuf, Hamda H Mohamed, Nimao A Ali, Zakhiya M Ahmed, Yousuf M Abdi

## Abstract

**Purpose:** this research aimed to identify the factors that lead to patients having mistrust towards the healthcare system in Somaliland and use the findings as a foundation for future policies.

**Methods:** a descriptive study was conducted to understand the factors contributing to the population’s mistrust of the healthcare system in Hargeisa. Data was collected through interviews with 49 participants using a survey questionnaire administered by interviewers, and convenience sampling was used to select participants based on availability and accessibility.

**Results:** through our analysis, we identified 11 themes that investigate issues influencing healthcare quality, cost, and accessibility, such as financial barriers, diagnostic accuracy, drug use, equipment quality, laboratory role, doctor knowledge and skills, medical ethics, governance, local doctor reputation, foreign healthcare, and the quality of available healthcare services.

**Conclusion:** patient -physician mistrust in Somaliland can be addressed by implementing laws, examining doctors’ knowledge, and examining hospital facilities. Improving public health sectors and limiting patient payments can reduce healthcare costs. Regulations on drugs can reduce medication errors and restore trust in the healthcare system.

## Introduction

### Statement of the Problem

Trust is an important part of the relationship between a patient and their doctor and one definition of trust states that trust is “The optimistic acceptance of a vulnerable situation in which the truster believes the trustee will care for the truster’s interests” ^1^. This definition shows that a patient expects that a healthcare provider will do their due diligence in helping them and not bringing harm to them because of their vulnerability.

Somaliland, a self -governing region in Somalia is located on the Horn of Africa and shares a coastline with Djibouti, and Puntland, which disputes some territorial claims. Somaliland’s GDP is around $2 billion, primarily remittances from Somalilanders working abroad. Its main export is livestock, shipped to neighboring Djibouti and Ethiopia, as well as Gulf states like Saudi Arabia and Oman. The country’s GDP per capita is one of the lowest in the world ^2^.

Boasting an estimated population of 4.3 Million mistrust in healthcare providers is one of the big challenges that face Somaliland’s healthcare system, in a country already dealing with a shortage of human resources, which remains one of the biggest challenges facing Somaliland utilization rate of public health facilities which remains very low ^3^.

## Background

Studies in foreign countries show similar trends of patients reporting mistrust against the healthcare establishment. Studies conducted on Chinese populations have shown significant mistrust in clinics ^4^, while at the same time studies conducted in the USA have shown that a significant portion of the respondents reported mistrust of certain parts of the US healthcare ^5^. Furthermore, studies in China, India, and Pakistan ^6–8^ have also shown evidence of many cases where mistrust towards the healthcare system, has led to violence of different types being carried out against healthcare workers.

## Purpose of the Study

Research on this topic in third -world nations like Somaliland is not widespread, thus this study was conducted to better highlight the biggest factors that lead patients in Hargeisa, to have a mistrust towards the healthcare system and perhaps become a foundation for further research into this topic, to hopefully find an effective solution to solve this problem.

## Material and methods

### Research Design

To answer our question a descriptive research design was agreed upon. A descriptive study is designed to describe the distribution of one or more variables, without regard to any causal or other hypothesis ^9^. This design is useful when the research aims to focus on gaining a deeper understanding of participants’ experiences and perceptions regarding a particular topic.

This research design helped us build a foundation on a topic that is not well understood enough in the context of Hargeisa and even the entirety of Somaliland.

### Sample for Data

The target population for the interviews was people who were not actively working as doctors to determine the factors that led to the population having a mistrust towards the healthcare system of Hargeisa.

The data was collected starting from **the 1st of June 2023 till June 14th** of the same year. 49 participants were interviewed, from 7 districts in Hargeisa - the capital of the self -declared country - Somaliland. These districts were as follows: 26 June, Ahmed Dhagah, Ahmed Moallim Haruun, Gacan Libaax, Ibrahim Kodbuur, Mohamed Moge and Mohamoud Haibe.

### Data Collection and Instruments

The Data was collected using a survey questionnaire that consisted of open -ended and closed - ended questions that were made and hosted on Google Forms. The questionnaires were administered by the interviewers personally instead of the participants. This allowed us to explain the questions to the participants in -depth and clarify any questions and doubts they had about the questions themselves^10^.

The questionnaires consisted of close -ended questions that asked for the participants for their demographic data such as: Gender, Age, Education level, Employment status and their jobs. Afterward, the participants were asked by the interviewers to give an answer on their experiences and the factors that led to them mistrusting the healthcare system of Hargeisa and largely Somaliland.

Convenience sampling a non -probability sampling technique in which participants are selected based on their availability and accessibility to the researcher, was used to interview the participants of this study ^11^. This method of sampling was used because it allowed us to interview enough people without the need for funding and investment of a considerable amount of time.

### Data Analysis

QDA Miner a software program developed by Provalis Research is a tool that allows researchers to organize, code, and analyze qualitative data in an efficient manner ^12^.

Using this software the answers from the questionnaire were coded. These codes were further analyzed using thematic analysis, defined as the process of identifying patterns or themes within qualitative data ^13^.

This helped us link the different responses given to us by the participants and consolidate the data we got into a set of defined ideas and themes, which further helped us understand what these results mean.

## Results

### Financial Problems

20 out of 49 have stated that they faced some sort of difficulties related to money when seeking medical care in Hargeisa. The high cost of healthcare was also one of the major issues reported by the participants. This included complaints about health services and medications being expensive, and the lack of discounts that can help patients offset the financial burden.

“High cost of medical care.” - Home teacher, Male.

“Lack of discounts.” - Teacher, Male.

One of the more worrying findings that we found through our interviews was some patients not being able to get medical services because they didn’t have money, and some of them even went further and explained that doctors put the pursuit of monetary gain above their sworn duty towards their patients.

“Without money, your needs are not attended to.” - Hotel -owner, Male.

“Some doctors, but not all, prioritize profit over patient care.” - Business owner, Female.

### Drugs and Prescription -Related Issues

21 out of 49 participants were basing their mistrust due to drug accompanying problems, and some of them apprised us that their health problems increased after they got the drug that some doctors prescribed to them.

Irrational drug prescription is one of the leading issues that all participants informed us about during data collection. Almost all participants condemn the poor handwriting of the doctors during prescribing medications which sometimes causes pharmacists to give some patients the wrong medication.

Lack of consent for the rational use of the drug, side effects of medications, and wrong dosing are some of the greatest challenges that every patient faces during visiting healthcare providers. Shortage of available drugs occurs sometimes, which makes some patients lose their trust in the health care system of the country.

Nonetheless, all our participants were worried about the quality of the drug, since the countries do not have quality control labs to check the standard of the drugs, the lack of effective quality control institutions gives chances some wholesalers to import the country low -graded drugs to make maximum profit from it. Our participants affirmed that this is one of the main reasons to travel to foreign countries to seek health care.

“He nearly killed me by administering an overdose of Hypertension medications “Retired midwife, Female.

“No drug quality” – Social worker, Male.

“They always prescribe with poor handwriting which harms patients.” - Cashier, Male.

### Knowledge and Skills of a Doctor

17 individuals out of 49, complained about the knowledge and skills of the doctors, citing that doctors don’t instruct the patients to return for a follow -up. They also don’t carry out a comprehensive history taking nor do they educate the patient well enough on their conditions, treatment plans, and future outcomes. They also mentioned that doctors don’t listen to patients, which can lead to misunderstandings between patients and doctors.

Participants also mentioned that our country lacks an adequate number of specialists leading to general practitioners treating complicated cases or doing high -risk surgeries, which puts patients’ lives at unnecessary risk.

“Not enough time is given to the patient’s history and examination” - Sales worker, Male.

“Doctors don’t have the adequate knowledge and some of them don’t have the right credentials” - Consultant, Male

“Our country doesn’t have many specialists, many of our medical professionals are general doctors -trying to cure every disease instead of a suitable specialist, and a lot of the diseases that are rare or complicated need a specialist, and some of the procedures can’t be made in our country” - Teacher, Female.

“Doctors not giving adequate information to patients leading to misunderstanding” - Housewife, Female.

### Misdiagnosis

Almost 14 out of 49 of our participants stated that both private and public health sectors carry out a lot of misdiagnoses, which can sometimes cause the loss of beloved family members due to an easily curable disease.

One of the participants clearly stated that she lost one of her family members due to a misdiagnosis which caused her to lose trust in the healthcare system.

Another participant told us that his daughter was taking pneumonia management for a whole week, but another doctor discovered that his daughter was suffering from acute viral hepatitis instead of pneumonia.

“The loss of one of my family members as a result of a misdiagnosis has left a deep wound in me” – Cashier, Female.

### Medical Equipment and Facilities

9 individuals out of 49 of our participants affirmed that there is: a lack of high -quality medical equipment, shortage of equipment, and even poor availability for more advanced medical equipment. A fair few contributors mentioned many incidents of failing medical equipment during procedures or when it is needed.

Numerous investigational types of equipment caused wrong results, which led to some patients traveling to foreign countries to seek more accurate and advanced medical equipment.

“Unavailability of advanced medical equipment” - Housewife, Female.

“No equipment.” - Accountant, Male.

“Lack of developed facilities and enough equipment to perform the various procedures.” - Student, Male

### Ethics and Behavior of a Doctor

13 of the 49 participants mentioned that the factors that contributed to their mistrust towards the doctors and largely Somaliland’s healthcare, were doctors not being attentive to their patients and prescribing medications and sending for medical tests with good intentions, but instead to make more money through unnecessary expenses on the patient’s part.

“When you come to the hospital doctors will send you to get unnecessary lab tests for money and profits this will be more difficult for the patients.” - Social worker, Male.

“Prescribe unnecessary medication for sale purposes.” - Consultant, Male.

“If you walk around some wards you can see many patients that are neglected with swollen hands from the cannulas” - Businessman, Male.

Participants also brought up that some doctors don’t have the behaviour and attitude expected of a doctor. They mentioned that doctors don’t devote time and attention to their patients; treat their patients with kindness and honesty; and professionalism; and practice good ethical practices in their work, and finally some participants noted that doctors are discriminatory or have some bias against some subset of the population.

“They are inhuman and don’t have a kind character.” - Laboratory technician, Male.

“No professional attitude.” - Unemployed, Male.

“They don’t listen and don’t pay attention to the patient.” - Home teacher, Female.

“Unethical workers.” - Teacher, Female.

“Lack of honesty.” - Librarian, Female.

“Some doctors have poor personalities.” - Government worker, Male.

“The doctors being Discriminatory and Biased.” - Manager, Male.

### Governance and Administration

A total of 3 participants of the 49 have expressed concerns with hospital administration and the government. They pointed out that qualified healthcare workers are not hired readily by hospitals. The lack of standard guidelines that are set by the relevant medical bodies in this country, was also one of the mentioned factors by the participants - that led to mistrust towards the healthcare system in Hargeisa and the rest of the country.

“The qualified health workers are not hired.” Teacher, Male.

Another factor that was mentioned by the participants, as one of the causes leading to mistrust against doctors and the healthcare system was - doctors not being held accountable for their actions, leading to patients not getting their rights and suffering because of that.

“There are no systemic regulations which control faults of medical workers, l saw a lot of people that were harmed due to medical errors - from severe complications to disabilities. I couldn’t also trust doctors because they would not agree to any talks or even admit their mistakes. This is caused by lack of clear guidelines which lead to them to the right path and help patients get their rights, and be compensated for any damage, but today patients are always the victims.” - Social Worker, Male.

### Reputation of Local Doctors

6 individuals mentioned that the local doctors don’t have the best reputations opting for foreign doctors citing a lack of trust. A participant also mentioned that media outlets also contribute to public mistrust.

“No trust between patient and doctor.” - Social Worker, Male.

“Misunderstanding between patients and doctors due to propaganda” - Saleswoman, Female.

### Foreign Healthcare and Doctors

4 participants mentioned the aspect of foreign healthcare being one of their main causes of mistrust, they reasoned that foreign countries have a better reputation due to them providing better healthcare. This can be because foreign countries have cheaper healthcare which leads to them being more trusted.

“I have had pain for several years in the abdomen and l went to different hospitals they told me that l have a disease in my stomach, and when l took my drugs l got no response. After many years l went to Ethiopia with my family members and they told me that l have a vascular disease, different than the previous one. They give me drugs and never felt again any abdominal discomforts” - Housewife, Female.

“When l want to go hospitals l prefer white doctors because they are better than our doctors and l feel more comfortable when l see them” - Housewife, Female.

“High cost of healthcare compared to neighbouring countries.” - Student, Male. “Misunderstanding between patients and doctors due to propaganda” - Saleswoman, Female.

### Laboratories

When we questioned the participants it became evident that they had a problem with labs, 4 participants mentioned that there was a shortage of labs and of those labs, they encountered inaccurate lab results which played a role in them not trusting the doctors.

“Lack of proper medical testing.” - Unemployed, Female.

“Labs are inaccurate sometimes, which makes me check labs from other clinicians.” - Librarian, Female.

“Misunderstanding between patients and doctors due to propaganda” - Saleswoman, Female.

### Quality of Healthcare Service

8 participants have expressed their concern with the quality of the available healthcare system, and they mentioned how poor its quality is.

“Poor health care services.” - Unemployed, Female.

“Poor quality of care” - Teacher, Female.

## Discussion

### Diagnostic Issues

Misdiagnosis is a significant issue in healthcare and the data suggests that there is a correlation between misdiagnosis and the level of trust. 28.6% of our participants have reported that misdiagnosis in both private and public health sectors; is one the factors that could lead to misunderstandings and loss of trust between patients and doctors, and as one participant pointed may even take the life of a loved one.

“The loss of one of my family members as a result of a misdiagnosis has left a deep wound in me” – Cashier, Female.

Addressing these issues is essential to ensure the safety and well -being of patients and healthcare providers.

Misdiagnoses may be sample -specific, single -cell -specific, or technique -specific. Many are independent of technique, sample type, or sample quantity in that they could occur owing to general system failures, e.g. human errors. Such system failures should be considered avoidable and should be the first to be eliminated. Examples of system failures include mislabeling and misidentification of labeled samples ^14^.

Research on this issue and its effects have shown that misdiagnosis and other diagnostic -related issues are prevalent in Western countries like the United States, where studies estimated that approximately 12 million US adults every year, or at least 1 in 20 US adults are affected by diagnostic -related issues ^15^.

Similar studies in the UK also estimated that 5.2% of deaths have a 50% or greater chance of being preventable. The main causes associated with preventable deaths are poor clinical monitoring 31.3%, diagnostic errors 29.7%, and inadequate drug or fluid management 21.1% ^16^. In conclusion, individuals’ and family members’ misdiagnosis experiences were associated with a reduction in trust in their current physician. Furthermore, these misdiagnosis experiences have been shown to have an additive effect by a study conducted in Japan ^17^.

Some ways we could improve the issue of misdiagnosis in the context of Somaliland - is to train doctors to be more attentive to their patients and their presenting symptoms, but also hire qualified professionals that can be used to build strong laboratory and radiology departments. Hiring technicians who have the knowledge and skills to operate and maintain medical equipment is also important. We can also improve it by double -checking for any mistakes and taking extra caution when diagnosing patients.

### Money and the Problems it Breeds

A great majority of our study participants have shared the sentiment that money -related issues are one of the greatest factors that lead to the loss of trust between patients and the healthcare system in Somaliland.

One of the money -related issues is that Somaliland has one of the lowest income rates in the world and this affects the population when they are getting healthcare. Due to the high cost of medical care, the general population doesn’t usually seek healthcare as frequently as they should because of how expensive it is.

When it comes to financial aid, the government has declared that they are committed to building a healthcare delivery system that is capable of providing healthcare services of high quality, accessible to everyone, equitable to all, and protecting people from falling into poverty due to out-of -pocket health expenditure^18^, but this policy is still far from being practical and is just a promise.

Another tragic and alarming issue that faces the population is that healthcare service in some hospitals and clinics health service is tied to having the patients pay large amounts of money upfront to receive medical attention.

These absurd scenarios don’t end there as some doctors due to the pursuit of profits, abandon their sworn duty not to do harm to their patients and strive to do what’s best for them - leading to patients shouldering the cost of unnecessary medications and laboratory tests disguised as necessary, to make money.

This trend of doctors picking and choosing medication has also been observed in Pakistan, where physicians will promote medications from pharmaceutical companies that reward with monetary gifts and suppress those companies that don’t pay a sum of money, regardless of the cost and efficacy of the medication ^19^.

Doctors in Somaliland have been criticized by the population for not upholding their duty of kindness towards their patients, increasing the rift between patients and the healthcare system of Somaliland.

This may even lead to worse healthcare outcomes for patients, as a study conducted in Finland noted that cancer patients have an increased risk of committing suicide ^20^, this highlights the need for physicians to not just treat illnesses but also provide mental and emotional support for their patients.

These findings show that doctors need to strive to practice kindness as an integral part of their medical practice and be more open toward their patients ^21^

To address these concerns, the relevant government bodies and hospital administrations, need to put in place policies and guidelines that standardize the health services and treatment that doctors and healthcare centers across the country use.

These policies also need to be enforced, and any parties that are found to have broken the terms they agreed upon and those who violated their patients’ rights - need held accountable and to be punished accordingly, to restore the faith of the people in the healthcare system of Somaliland. Doctors also need to be educated on the cost of medications as a study found that Doctors tended to overestimate the cost of inexpensive drugs and underestimate the cost of expensive ones ^22^, demonstrating a need for easily accessible databases that contain medication costs, and also providing better training for doctors in this subject.

### Knowledge and Skills

In the eyes of the patients, doctors in Somaliland have a bad reputation, which can be attributed to the fact that a lot of patients and their families have undergone the experience of a doctor with inadequate knowledge and skills working for them, and this has caused them to lose trust in the doctors.

17 individuals out of the 49 we interviewed complained about the knowledge and skills of the doctors. They complained that doctors don’t instruct patients on follow -ups, don’t educate them on conditions, treatment plans, and future outcomes, and don’t listen to patients. This all comes under communication, and it can all be attributed either to a lack of knowledge in this respective field or to the lack of skills needed to implement such knowledge into a doctor’s routine. An interesting outlier of the answers we got was that the lack of specialists in the country led to general practitioners treating complicated cases or high -risk surgeries, putting patients’ lives at risk.

Recent data has shown that the standard of doctors, be it in terms of what is needed from them or how they do their day -to -day work, has improved. Current doctors are at all times required to follow the international SOPS, and a lot of those doctors graduated from universities that grade them using internationally recognized grading systems. A lot of them have adequate knowledge, but still, the stigma the public has against doctors keeps on going.

This isn’t a problem exclusive to us, as other research or literature reviews also highlight the importance of knowledge and skills and the effect a lack of them has.

Effective doctor -patient communication is a central clinical function in building a therapeutic doctor -patient relationship, which is the heart and art of medicine. This is important for the delivery of high -quality health care. Much patient dissatisfaction and many complaints are due to breakdowns in the doctor -patient relationship ^23^.

“When I started working in the NHS, I noticed that there was a major emphasis placed on communication skills in addition to clinical skills, which could be the first major difference between practicing medicine in the United Kingdom and in the Middle East where clinical skills are the primary focus in medical practice” ^24^.

Doctor -patient communication is a fundamental component of clinical practice. In addition to being knowledgeable scientific experts in various specialties, effective doctor -patient communication is required for building a therapeutic doctor -patient relationship. In recent years, a growing emphasis on patient autonomy, patient -centered care, and consumerism in medicine has further exemplified the importance of effective doctor -patient communication. Despite this, good doctor -patient communication remains a challenge for physicians and is the underlying reason for the majority of patient complaints ^25^.

A breakdown in the doctor -patient relationship often manifests as unsatisfactory patient -doctor communication, the dominant theme in malpractice claims. This could be related to the contrasting perspectives of patients and doctors on what constitutes effective communication. Patients prefer a psychosocial model of communication compared to a biomedical model, which is used more commonly by doctors. Doctors also tend to overestimate their communication abilities ^25^.

To improve trust and knowledge, healthcare providers should focus on providing adequate information and training to patients, ensuring that they receive the necessary care and support.

### Drugs and Prescription -Related Issues

Our study found that patients’ perceptions of drugs related to the quality of drugs, irrational prescription, wrong dosing of medications, lack of consent about sensible use of drugs, and the side effects of the drug contributed to patient -physician mistrust. From the patient’s perspective, illegible handwriting can delay treatment and lead to unnecessary tests and inappropriate doses which, in turn, can result in discomfort and death Poor handwriting remains a significant problem in medicine not only in Somaliland but this kind of problem was seen in many places of the world, this is due to in past centuries, doctors scribbled notes to keep a personal record of the patient’s medical history. The notes were generally seen only by the doctor. Today, doctors are no longer one -man bands. With dozens of other professionals, doctors are but one element of a large, multidisciplinary healthcare team. A consequence of this expansion is that illegible scrawls, hurriedly composed by rushed doctors, are now presented to colleagues with no qualifications in cryptology ^26^.

Illegible handwriting in medical records can have adverse medico -legal implications, some researchers in the US note that ‘few admissions look more damaging in testimony than physicians admitting they cannot read their hand -writing. There was a case where a physician’s poor penmanship led to a fatal overdose of an elderly patient. The physician was sued for $380,000 ^27^.

Many incidents were seen in developed countries, but every country has it is own drug regulation laws that protect patients from this kind of negligence and even compensate if it occurs, for example in America, the Medical Defense Union’s Ten Commandments of record keeping, **‘Thou shalt write legibly’** comes top of the list ^28^.

How to fix this problem? Making rules and regulations against this kind of malpractice, Handwriting tests as part of hospital appointments, and a sophisticated IT system to computerize patient notes.

Medical errors also occur in developed countries, The goal of medication use is to achieve defined therapeutic outcomes with improvement of quality of life and minimize patient risk ^29^. Medical errors can occur at any phase of the medication use cycle from prescribing, dispensing, and administration of a drug to the patient. It increases morbidity and mortality of the population along with an increase in the cost of the treatment. Further, it also affects patients’ confidence in medical care ^30,31^ we advise Probably, computerizing the medication process system in hospital settings and pharmacological education of prescribers and nurses could help to reduce medical errors.

Poor quality medicines have a large impact on the developing nation like Somaliland. Considering the vast scale of the global pharmaceutical industry and the incidence of potentially fatal diseases, any amount of poor -quality medicine is unacceptable because it increases morbidity and mortality. The impact of poor -quality medicines is most clearly evident if they contain lethal incorrect active ingredients ^32^ we suggest that the government bring good and effective labs to protect people from poor -quality drugs from pharmacist companies and to make rules and regulations about this issue.

## Conclusion

Patients in Hargeisa and generally in Somaliland express that they have lost trust in the doctors and the healthcare system. Through our research, we found that the causes of this loss of trust according to our participants were financially related problems that led to patients not being able to afford care, or doctors acting unethical practices in the pursuit of money. Other findings were a lack of high -quality and advanced equipment, facilities, and labs; an uncontrolled drug market, and widespread practice of irrational prescription of drugs.

We would recommend for these problems to be solved easily that the government makes laws and regulations against negligence and malpractice. The relevant Authorities should also put in place systems that check every doctor’s knowledge and skills before approval.

Ministry of Health must examine every hospital equipment and facility before giving them a license. Improving and developing effective public health sectors will reduce the amount of money that citizens spend on health care, also executive can limit the amount of money that patients pay for certain health services in private health sectors, evaluating and estimating the quality of caregiving is essential for transitioning away from ingrained profit -focused models.

Establishing advanced labs to protect drugs that enter that country improves the quality of the drugs. Probably, computerizing the medication process system in hospital settings and pharmacological education of prescribers and nurses could help to reduce Medication errors.

In addition, a drug use policy should be implemented and maintained to reduce inappropriate use of drugs. Trust -building training must be an explicit part of medical education, reforming medical education and promoting caregiving are pathways towards restoring trust. In addition to administrative and legal changes, a moral response is urgently needed to restore trust, for example, the public can be urged to report any negligence or malpractice by healthcare workers.

Areas that we would recommend for future research are the impact of patient mistrust of the healthcare system on their health outcomes; the role of media in patient mistrust and the solutions other countries used to address this issue.

## Supporting information

Survey questionnaire form

Survey responses

## Data Availability

All data produced in the present study are available upon reasonable request to the corresponding author

## Acknowledgments

First and foremost, we would like to acknowledge our supervisor, Dr. Zakiyah Jama, for taking part in reviewing our research. We would also like to acknowledge Mr. Mohamed Mussa for assisting us in developing the research outline and our methodology. We would also like to thank all our authors for their hard work and devotion towards completing this project. Lastly, we would like to thank God for enabling us to pursue this opportunity with good health.

## Disclosure

### Ethical Considerations

To make sure our study was ethical, this study was undertaken after gaining the College of Medicine and Health Science at the University of Hargeisa. The interviewers explained to the participants the kind of questions they would be asked and how that information was going to be used and only proceeded forward after gaining consent.

