## Supplementary material for "Factors that lead to patient mistrust of the healthcare system in Hargeisa, Somaliland: a qualitative exploration": Survey questionnaire form

### Research survey

This survey is part of research being conducted by medical students of the University of Hargeisa.

* Indicates required question

1. Email *

2. Informed consent *

   By submitting this survey you give consent for your responses to be used in this study, without identifying you personally and you affirm that you are at least 16 years of age. Do you consent?

   *Mark only one oval.*

   ( ) Yes

   ( ) No

Personal data

3. Area of residence

4. **Gender** *

   *Mark only one oval.*

   ( ) Male

   ( ) Female

5. **Age** *

*Mark only one oval.*

- 16 - 26
- 26 - 36
- 36 - 46
- 46 - 56
- 57+

6. **Education level** *

*Mark only one oval.*

- No formal education
- Primary school
- Intermediate
- Secondary
- University

7. If you are a university graduate, what was your university degree?

8. **Are you currently employed?** *

*Mark only one oval.*

- Yes
- No

9. **Occupation** *

Questions

10. What difficulties have you faced from doctors that made you have distrust of them? *

This content is neither created nor endorsed by Google.

Google Forms
